# Machine Learning and Artificial Intelligence in Health Services and Policy Research literature for Primary Health Care: A Scoping Review protocol

**DOI:** 10.1101/2025.08.01.25332615

**Authors:** Pablo Galvez-Hernandez, Li-Anne Audet, Zahra Shakeri, Walter P Wodchis

## Abstract

**Objective:** Artificial intelligence (AI) and machine learning (ML) are widely used in healthcare, primarily for clinical tasks like diagnostics and decision support. However, their role in organization- and system-level processes, such as resource allocation and workforce planning, remains underexplored. This scoping review aims to review AI and ML applications at the meso- and macro-levels of primary health care (PHC) systems reported in Health Services and Policy Research literature, assessing their strengths, limitations, and gaps to guide future research.

**Methods:** This scoping review will follow Arksey and O’Malley’s five-stage framework and PRISMA-ScR guidelines. A comprehensive literature search will be conducted in Medline, CINAHL, Embase, Cochrane Library, PsycINFO, and IEEE Xplore, as well as grey literature from OpenGrey, Google Scholar, and ProQuest. The search will cover January 2010 to December 2024, with a final search update in 2025 prior to manuscript submission to ensure inclusion of the most current evidence. Two independent reviewers will screen titles, abstracts, and full texts, resolving discrepancies by consensus. Eligible studies will include primary research describing, evaluating, implementing, or developing AI and ML applications at the meso-level (e.g., organizational monitoring and evaluation) and macro-level (e.g., system-wide funding and workforce/resource allocation) in PHC. Studies on micro-level applications, non-implemented research, and secondary literature will be excluded. Data will be extracted using a protocol and synthesized based on meso- and macro-level PHC dimensions, adapted from WHO’s operational and measurement frameworks. The protocol has been registered in Open Science Framework (OSF) (osf.io/wzj5x).

**Conclusions:** This review will synthesize AI and ML applications in organizational and structural dimensions of PHC, highlighting understudied areas and informing future research and policy. The findings will provide insights into AI and ML’s strengths and limitations in supporting critical PHC elements, such as governance, resource allocation and workforce planning.

## Background

Applications of Artificial Intelligence (AI) and Machine Learning (ML) in healthcare have grown exponentially over the past decade [1]. AI can be defined as a broad field focused on systems that exhibit intelligent behavior by deriving knowledge from data [2]. Machine learning (ML), commonly considered a subfield of AI, uses algorithms to learn associations with predictive power from data examples through statistical models [2]. These tools enable the analysis of large, complex data to uncover patterns and predict outcomes, offering promising advancements in how healthcare is delivered.

Most studies on AI and ML in healthcare have focused on their clinical applications, including diagnostic tools, chatbots, and predictive analytics embedded in healthcare IT infrastructure for patient care delivery [3, 4]. Additionally, there is increasing interest in population-based applications in public health, often referred to as precision population health management, which uses advanced analytics (e.g., AI, machine learning, and predictive modeling) to stratify populations and tailor interventions based on clinical, behavioral, and social data, aiming to deliver targeted, equitable care at scale [5]. Numerous empirical data have shown that AI and ML can assist clinicians to make better decisions, early diagnosis and patient monitoring [6, 7]. These benefits have also been synthesized in systematic reviews, which highlighted the advantages of AI and ML for patients, families, and healthcare professionals in various healthcare contexts [8, 9].

While these applications highlight the central role of healthcare delivery and clinical practice, effective healthcare systems also require well-organized and efficient underlying processes and structures that support patient care. Elements such as policies, funding, governance, workforce planning, leadership, and organization of care pathways play a critical role in enabling care delivery that addresses population needs [10]. These meso-level (organizational and team-level) and macro-level (regional and national political, regulatory, and funding structures) factors underpin the foundation of healthcare systems as well as the day-to-day practice of interprofessional teams and clinicians [11]. However, the potential of AI and ML applications to strengthen and understand these structural and systemic elements remains largely unexplored, representing a significant gap in current research.

Within health systems, Primary Health Care (PHC), a cornerstone of efficient health systems and universal health coverage, faces persistent challenges in sustaining high-quality service delivery [12]. Examples of structural and process-related challenges include resource allocation favouring specialized care; difficulties in planning and funding PHC strategies that involve multisectoral collaboration; adapting care models to local contexts; inadequate workforce planning amid shortages; inefficiencies in payment and purchasing systems; and deploying integrated monitoring and evaluation frameworks [12, 13]. These challenges are compounded by an aging population with complex multimorbidity and growing social needs, placing additional strain on PHC systems [14]. While economic and workforce resource constraints present a major burden, existing infrastructures could be strengthened through more effective resource allocation, streamlined care pathways, needs-based service and workforce planning, and better integration supported by timely data [15, 16]. AI and ML might have the potential to support these improvements; however, their role in addressing these challenges remains unclear and has not been comprehensively synthesized in existing reviews.

Some empirical research has explored the macro- and meso-level applications of AI and ML in primary health care systems. For instance, Orlando et al. used AI-driven clinical decision support tools for precision population health, enabling risk stratification in primary care populations and enhancing care delivery across diverse primary care settings [17]. Similarly, Rajkomar et al. applied unsupervised learning techniques, including k-means clustering combined with log-linear regression models on electronic health record data, to identify patient groups phenotypes, predict future healthcare utilization, and develop weighted primary care panel sizes to optimize provider workload distribution [18]. Recent investigations have begun to apply machine-learning techniques to administrative challenges, such as optimising appointment scheduling, although this line of work remains in its nascent stage [19]. However, a comprehensive understanding of AI and ML applications in the broader structural and organizational dimensions of primary care is missing, as existing reviews have primarily focused on micro-level applications, those involving individuals, such as patients and providers, and their interactions. For example, a 2021 scoping review by Abbasgholizadeh et al. found that diagnosis, risk stratification, disease surveillance, and clinical decision support using mainly machine learning and natural language processing were the primary applications of AI and ML in PHC [8]. AI techniques are beginning to be applied to these structural challenges, for example to support strategic purchasing or funding allocation decisions in health systems, although evidence within the PHC context remains limited [20].

Addressing the critical gap in examining AI and ML applications and their strengths and limitations for system and organizational-level dimensions of PHC is essential for understanding how these technologies could impact core elements including health system governance resource allocation, workforce planning, and intersectoral coordination. As Health Services and Policy Research is the field of research that focuses on studying the structural and organizational elements underpinning health care systems, this scoping review aims to (i) map the current applications of AI and ML in meso and macro levels of PHC within Health Services and Policy Research literature, (ii) assess the strengths and limitations of these tools, and (iii) identify application gaps and future directions in the field.

## Methods

This scoping review will follow the five stages proposed by Arksey and O’Malley: (i) identifying the research questions; (ii) identifying relevant studies; (iii) study selection; (iv) charting the data, and (v) collating, summarizing and reporting the results [21]. Results will be reported according to the PRISMA Extension for Scoping Reviews (PRISMA-ScR) checklist [22]. This scoping review was registered in Open Science Framework (OSF) (osf.io/wzj5x). Ethics approval was not required for this study because human subjects are not involved.

### Scoping review stages

#### Stage 1: Defining the research questions

The review will be guided by the following research questions: (i) What are the current applications of machine learning (ML) and artificial intelligence (AI) in the Health Services and Policy Research literature in meso and macro elements of primary health care? (ii) What are the strengths and limitations of these applications in addressing meso and macro-level challenges within PHC systems?

#### Stage 2: Identifying relevant studies

Studies will be searched in six databases: Medline, CINAHL, Embase, Cochrane Library, PsycINFO, and IEEE Explore. In addition, a grey literature search will be conducted in Opengrey, Google Scholar and Proquest. Finally, reference lists of included articles and relevant literature reviews will be searched to ensure all relevant studies had been included [21]. The search will be restricted to publications in English between Jan 2010 and Dec 2024, with a final search update conducted in 2025 prior to manuscript submission, to ensure the review reflects the most current evidence available at the time of publication. The start date was chosen to capture the period of rapid expansion of AI and ML in healthcare, marked by the widespread adoption of techniques such as deep learning algorithms in the early 2010s [23].

A search strategy was iteratively developed following the recommendations of the Joanna Briggs Institute Reviews Manual [24]. Initially, a limited search was conducted in Medline OVID using the key terms “artificial intelligence”, “machine learning”, and “primary health care”, identifying a sample of five relevant articles. This was followed by an analysis of the index terms and keywords using the Yale Mesh Term Analyzer [25]. Subsequently, a comprehensive search strategy was developed, incorporating identified keywords and index terms, combined with Boolean operators to capture two main concepts: AI and ML, and Primary Health Care. The final search strategy (shown in Table 1) was reviewed for sensitivity during a consultation with an expert librarian at the University of Toronto. The strategy was then tailored for specific lexicon across databases. As part of the search validation procedure, five hand-searched articles will be used to validate the final search strategy and ensure its effectiveness in capturing relevant studies. We chose not to include ‘data mining’ as a separate term because it overlaps heavily with machine learning and was less specific, but we remained alert to such terminology during screening.

**Table 1.**
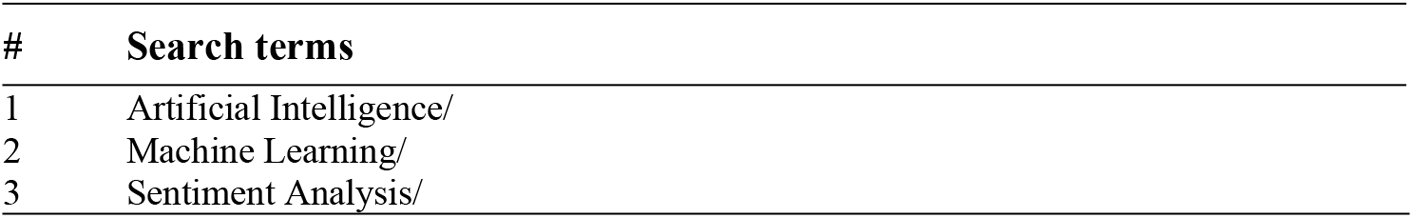
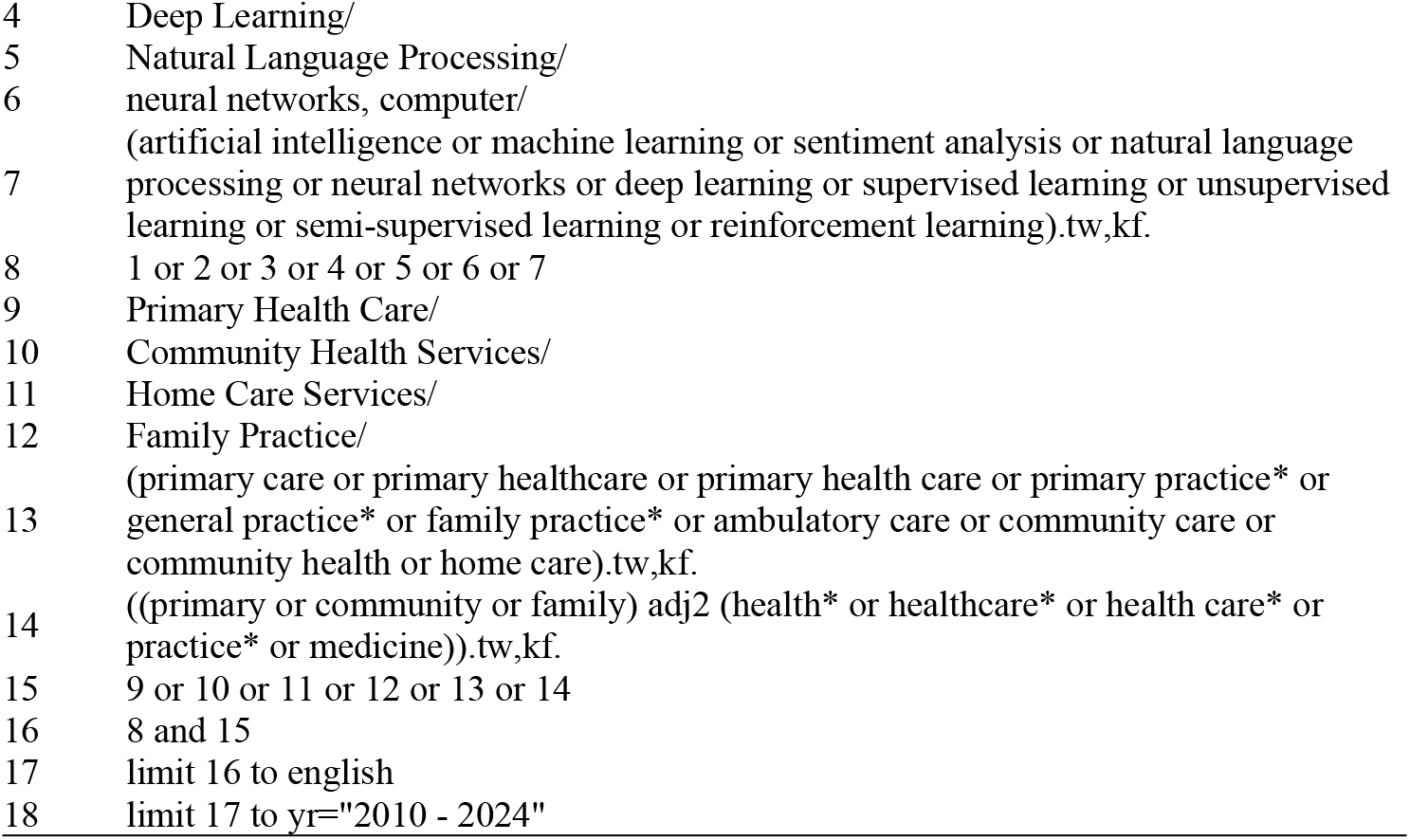
Search strategy for Medline OVID.

#### Stage 3: Study selection

All references will be uploaded to Rayyan software for deduplication and screening. Two independent reviewers (PGH, LA) will conduct the screening process in three stages, title, abstract, and full text, using detailed protocol outlining the eligibility criteria (S1 File). To ensure rigor, we will first validate the screening protocol by conducting a training session in which two reviewers will simultaneously screen a sample of 25 articles [21]. Two reviewers will subsequently conduct a calibration exercise by independently screening a test set of 20% of articles at each stage (i.e., title, abstract and full text) with a targeted inter-rater reliability of over 70% [26]. Discrepancies will be resolved in periodic follow-up meetings to discuss decision rationales, address inconsistencies, and reach consensus with the involvement of a tie-breaker when necessary.

The eligibility criteria are defined a priori, drawing from previously established conceptual definitions included in Table 2. *Inclusion criteria* are as follows: (i) published and unpublished primary studies in the field of Health Systems and Policy Research that (ii) describe, evaluate, implement, or develop AI and ML methods (iii) at the meso and macro levels of Primary Health Care (PHC) systems. For this review, AI methods primarily refer to computational techniques that enable learning or data-driven decision-making (machine learning). Knowledge-driven AI (e.g., expert systems) would be included if they appear in the context of PHC systems, but our preliminary search suggests most applications are ML-based. By focusing on meso/macro levels, we target studies where AI/ML is applied to organizational processes or system challenges (e.g., AI used for clinic management, health workforce planning, primary care policy analysis, or resource allocation across services). Studies purely addressing individual patient care (even if using AI) are beyond our scope.

**Table 2.** Conceptual definitions underpinning inclusion and exclusion criteria. These categories are not mutually exclusive (e.g., deep learning methods are frequently applied in NLP tasks), but they provide a helpful framework for classifying AI applications by their primary technique and data type).

| Concept | Categories and definitions |
| --- | --- |
| Artificial intelligence techniques | AI techniques and their definitions are broadly classified into three groups [27]:<br>*Traditional machine learning (ML)*: Algorithms used to classify and predict structured data.<br>*Advanced deep learning*: Techniques such as convolutional neural networks to analyze high-dimensional patterns in large, complex datasets like medical images.<br>*Natural language processing (NLP)*: Techniques to analyze unstructured text and handwritten data. |
| Machine learning algorithms | ML algorithms are categorized into four groups based on learning approaches [27, 28]:<br>*Supervised learning:* Algorithms that are trained on labeled data to map inputs to outputs, supporting tasks such as classification and regression [27, 28].<br>*Unsupervised learning:* Algorithms that analyze unlabeled data to uncover hidden patterns or structures in those data.<br>*Semi-supervised learning:* Algorithms that train on a small amount of labeled data combined with a larger pool of unlabeled data, effectively combining supervised and unsupervised approaches (e.g., using a few labeled patient records to inform learning from many unlabeled records).<br>*Reinforcement learning* involves algorithms learning through interaction with an environment, using feedback (rewards or penalties) to optimize actions. |
| AI and ML methods | The specific methods underlying the AI and/or ML application, such as deep neural networks, support vector machines, decision trees, k-nearest neighbors, and random forests [2, 28]. |
| Primary Health Care and Primary Care | *Primary health care (PHC):* A comprehensive, society-wide strategy aimed at organizing and enhancing national health systems to deliver health and well-being services closer to communities [29].<br>*Primary care:* The settings that constitute the initial point of contact with the healthcare system, such as primary care centers, community health centers, and community pharmacies [29]. |
| Meso and macro levels of PHC systems | *Meso level:* Refers to the organizational or local institutional level of PHC systems, including elements such as organizational dynamics, team-level processes and structural elements like facility budgets and local intersectoral partnerships [30].<br>*Macro level:* Encompasses broader influences, such as national and provincial policies, scope of practice regulations, and national budgeting [30]. |
| Health Services and Policy Research | A multidisciplinary field of inquiry, both basic and applied, that investigates how social factors, financing systems, organizational structures and processes, health technologies, and personal behaviors affect the delivery, quality, access, cost, and outcomes of primary health care, as well as the development and impact of policies [11]. |

All categories and techniques of AI and ML will be included (e.g., supervised, unsupervised, semi-supervised, reinforcement, deep learning; natural language processing, etc.). Unpublished primary studies will include dissertations, theses, conference papers and proceedings, and reports.

*Exclusion criteria* include: (i) non-implemented research (e.g., commentaries, editorials); (ii) secondary studies, such as literature syntheses (iii) studies in PHC reporting micro-level applications of AI and ML (e.g., digital technologies for patient care, diagnostic tools); (iv) studies conducted in settings other than PHC systems; and (v) non Health Services and Policy Research literature, such as clinical, public health, or epidemiological studies focusing solely on efficacy of technologies on patients or measuring population health profiles and patterns. We will include studies regardless of the venue (clinical or technical journals are not excluded) as long as the study’s focus is on meso/macro-level PHC system applications. Studies that only address individual clinical predictions without organizational/system context will be excluded as micro-level.

#### Stage 4 & 5: Charting the data, collating, summarizing and reporting the results

The data charting process will involve a training phase for extractors, a reliability check, and the final extraction stage. A pilot test will be conducted on a subset of 10% of included studies to refine the charting process prior to full data extraction. One reviewer (PGH) will chart the data, which will be independently verified by a second reviewer (LA). Any discrepancies will be resolved through discussion or by a third reviewer if necessary.

The following information will be charted for each included study: (i) *Descriptive study information* including title, author(s), year of publication, country/region, journal/source, study type, study objectives, funding source; (ii) *AI and ML applications:* a brief description of the application’s purpose, AI/ML technique categories (traditional ML, deep learning, NLP), type of ML approach (e.g., supervised, unsupervised, semi-supervised, or reinforcement learning), and specific algorithms used (e.g., decision trees, convolutional neural network, k-nearest neighbors); *(iii) Primary health care system data*: PHC system level (meso, macro), type of setting (e.g., primary care centers, community health centers), target population, PHC system dimension (structure, process, outcome), PHC area of focus (e.g., governance, funding, service planning, workforce planning); and (iv) *main findings and strengths and weaknesses of the AI/ML tools, as identified* by the authors.

The process of collation will involve an initial step of coding and categorizing the extracted data using labels by the categories described in Table 2, such as type of AI techniques and algorithms employed. Subsequently, AI and ML applications will be collated and mapped based on the specific meso- and macro-level areas of PHC targeted. For this purpose, we have defined a list of PHC meso- and macro-level areas (Table 3), categorized into structures, processes, and outcomes, adapted from two WHO Operational and Measurement frameworks for Primary Health Care [12, 15]. These frameworks identify levers to accelerate progress in strengthening PHC, that include core elements such as funding and allocation or resources, engagement of community and other stakeholders, health care workforce and purchasing and payment systems.

**Table 3.** Meso and macro elements of PHC systems from a HSPR perspective.

| PHC areas | Level |
| --- | --- |
| **Structures**<br>Political commitment and leadership<br>Governance & policy frameworks<br>Community and stakeholder engagement structures<br>Monitoring and evaluation<br>Funding and allocation of resources<br>Availability of PHC facilities and infrastructure for communication and transportation<br>PHC workforce density, distribution, accreditation and education<br>Medicines and health products<br>EMR, surveillance and digital health systems | Macro<br>Macro/Meso<br>Meso<br>Macro/Meso<br>Macro/Meso<br>Meso<br>Macro/Meso<br>Macro/Meso<br>Meso |
| **Processes**<br>Selection and planning of services and estimation of demand<br>Service design: Empanelment systems, patient referrals and care pathways for tracer conditions.<br>Organization and facility management capability and leadership, team-based service delivery and facility budgets<br>Community engagement for intersectoral collaboration, planning and organization, and proactive population outreach<br>Systems for quality improvement: access, effectiveness, safety and efficiency. | Macro/Meso<br>Meso<br>Meso<br>Meso<br>Macro/Meso |
| **Outcomes**<br>Population-based equity, access, affordability, acceptability, availability and utilization of PHC services.<br>Population-based quality of care, effectiveness, safety, efficiency, and costs. | Macro/Meso<br>Macro/Meso |

Results will be synthesized thematically according to the PHC elements shown in Table 3. To explore whether different methodological approaches are applied to different PHC challenges, findings will also be reported stratified by AI technique category and learning type. The results will be presented narratively, supported by charts and tables for visualization. The strengths and limitations of AI/ML tools, as reported by the study authors, will be compiled, categorized (e.g., by AI application or PHC element), and analyzed separately. The review process will be detailed using the PRISMA Extension for Scoping Reviews (PRISMA-ScR) flow diagram to ensure transparency, as well as the PRISMA-P checklist (S2 File). Because this scoping review aims to synthesize existing research and provide a comprehensive overview of AI applications in PHC, rather than focusing on study outcomes, a risk of bias assessment will not be conducted.

## Discussion

This scoping review protocol outlines a systematic approach to mapping AI and ML applications across meso- and macro-level dimensions of primary health care systems. The review has completed Stage 3 (study selection). Completion of the full scoping review is expected by December 2025. By synthesizing evidence from published and unpublished literature across six major databases and grey literature, the review will provide insights into how AI and ML have been applied to support organizational processes, governance, resource allocation, workforce planning, and intersectoral coordination in PHC settings. This focus extends beyond the predominantly micro-level clinical applications explored in previous reviews, addressing a gap in health services and policy research. To ensure methodological rigor and feasibility within resource constraints, this protocol focuses on peer-reviewed and grey literature in English and utilizes specific “primary health care” terminology, which might limit the inclusion of research from regions where first-level health care services may use differing nomenclature.

Despite these limitations, this review will provide a comprehensive overview of how AI and ML technologies are applied at understudied levels of PHC, where their use has not yet been synthesized. The findings will contribute to Health Services and Policy Research in PHC by identifying gaps and emerging AI and ML applications, as well as informing recommendations to strengthen the structural and organizational capacity of PHC systems.

## Data Availability

All relevant data are within the manuscript and its Supporting Information files.

## Supporting information

S1 File. Screening protocol

S2 File. PRISMA-P Checklist

## Acknowledgments

The authors would like to thank the Faculty Liaison & Instruction Librarian at University of Toronto for their input when designing and validating the search strategy.

## Author Contributions

**Conceptualization:** Pablo Galvez Hernandez.

**Methodology:** Pablo Galvez-Hernandez, Li-Anne Audet, Zahra Shakeri, Walter P Wodchis.

**Supervision:** Walter P Wodchis, Zahra Shakeri.

**Writing – original draft:** Pablo Galvez-Hernandez.

**Writing – review & editing:** Pablo Galvez-Hernandez, Li-Anne Audet, Zahra Shakeri, Walter P Wodchis.

**Guarantor of the review:** Pablo Galvez Hernandez.

**Competing interests** The authors declare no conflicts of interest.

**Patient consent for publication** Not required.

